# Computational Deconvolution of Cell Type-Specific Gene Expression in COPD and IPF Lungs Reveals Disease Severity Associations

**DOI:** 10.1101/2024.03.26.24304775

**Authors:** Min Hyung Ryu, Jeong H. Yun, Kangjin Kim, Michele Gentili, Auyon Ghosh, Frank Sciurba, Lucas Barwick, Andrew Limper, Gerard Criner, Kevin K. Brown, Robert Wise, Fernando J. Martinez, Kevin R. Flaherty, Michael H. Cho, Peter J. Castaldi, Dawn L. DeMeo, Edwin K. Silverman, Craig P. Hersh, Jarrett D. Morrow

## Abstract

**Rationale:** Chronic obstructive pulmonary disease (COPD) and idiopathic pulmonary fibrosis (IPF) are debilitating diseases associated with divergent histopathological changes in the lungs. At present, due to cost and technical limitations, profiling cell types is not practical in large epidemiology cohorts (n>1000). Here, we used computational deconvolution to identify cell types in COPD and IPF lungs whose abundances and cell type-specific gene expression are associated with disease diagnosis and severity.

**Methods:** We analyzed lung tissue RNA-seq data from 1026 subjects (COPD, n=465; IPF, n=213; control, n=348) from the Lung Tissue Research Consortium. We performed RNA-seq deconvolution, querying thirty-eight discrete cell-type varieties in the lungs. We tested whether deconvoluted cell-type abundance and cell type-specific gene expression were associated with disease severity.

**Results:** The abundance score of twenty cell types significantly differed between IPF and control lungs. In IPF subjects, eleven and nine cell types were significantly associated with forced vital capacity (FVC) and diffusing capacity for carbon monoxide (D_L_CO), respectively. Aberrant basaloid cells, a rare cells found in fibrotic lungs, were associated with worse FVC and D_L_CO in IPF subjects, indicating that this aberrant epithelial population increased with disease severity. Alveolar type 1 and vascular endothelial (VE) capillary A were decreased in COPD lungs compared to controls. An increase in macrophages and classical monocytes was associated with lower D_L_CO in IPF and COPD subjects. In both diseases, lower non-classical monocytes and VE capillary A cells were associated with increased disease severity. Alveolar type 2 cells and alveolar macrophages had the highest number of genes with cell type-specific differential expression by disease severity in COPD and IPF. In IPF, genes implicated in the pathogenesis of IPF, such as matrix metallopeptidase 7, growth differentiation factor 15, and eph receptor B2, were associated with disease severity in a cell type-specific manner.

**Conclusion:** Utilization of RNA-seq deconvolution enabled us to pinpoint cell types present in the lungs that are associated with the severity of COPD and IPF. This knowledge offers valuable insight into the alterations within tissues in more advanced illness, ultimately providing a better understanding of the underlying pathological processes that drive disease progression.

## INTRODUCTION

Chronic obstructive pulmonary disease (COPD) and idiopathic pulmonary fibrosis (IPF) are debilitating chronic diseases of the lungs with progressive and complex pathobiology [1,2]. COPD is characterized by airflow limitation, chronic airway inflammation, and lung parenchymal destruction [1]. IPF is characterized by cellular proliferation, interstitial inflammation, and fibrosis [2]. COPD and IPF are both related to long-term inhalation of noxious agents (e.g. tobacco smoking) and manifest in older adults as accelerated lung aging [3]. As such, both diseases are associated with significant morbidity, mortality, and a high economic burden to our society [4,5]. Therefore, there is an urgent need for disease prevention and improved treatments.

Genetics plays a role in predisposition to both diseases; eighty-two and nineteen loci have been associated with the risk of developing COPD or IPF, respectively [6,7]. COPD and IPF risk loci are enriched for pathways important in regulating cellular functions. For example, COPD risk loci are enriched for pathways regulating extracellular-matrix, cell-matrix adhesion, histone deacetylase binding, the Wnt-receptor signaling pathway, SMAD binding, and the MAPK cascade [6]. Similarly, IPF risk loci are enriched for pathways related to host defense, cell-cell adhesion, spindle assembly, transforming growth factor beta (TGF-β) signaling regulation, and telomere maintenance [8]. Furthermore, genetic factors are postulated to impact disease susceptibility in a cell type-specific and context specific manner. Therefore, improved molecular characterization of cells in the diseased lungs may provide insight into understanding disease pathobiology, paving the path to new therapeutics.

Investigating the molecular and cellular aspects of pathological lungs in the context of these diseases holds great promise for developing preventative and treatment strategies. In particular, single-cell RNA sequencing (scRNA-seq) has been used in COPD and IPF patients to search for putative disease-causing cell types. For example, scRNA-seq analysis of IPF lungs has identified aberrant basaloid cells, a rare, disease-enriched cell type [9]. In COPD lungs, scRNA-seq has identified a high metallothionein-expressing macrophage subpopulation enriched in advanced COPD and altered bioenergetics and cellular stress tolerance in an alveolar type 2 pneumocyte (ATII) subpopulation [10]. A recent multi-omic single-cell analysis revealed a CD8^+^ T cell subpopulation (KLRG1+TEMRA cells) to be enriched in COPD lung tissue [11]. However, the number of subjects included in these prior studies was modest, limiting the generalization to a larger patient population.

Due to the cost and technical limitations, performing scRNA-seq or tissue dissection experiments combined with fluorescence-activated cell sorting are yet to be practical in large epidemiology cohorts (n>1000). Moreover, the impact of tissue dissociation on gene expression in fluorescence-activated cell sorting (FACS) and scRNA-seq protocols remains poorly understood. Given that COPD and IPF are heterogeneous diseases, molecular studies encompassing a wide range of subjects with cell type-specific resolution are needed to unravel the complex interplay of cells in disease pathophysiology. To this end, large-scale clinical and genomic data in population cohorts may be leveraged to advance our search for cellular drivers of COPD and IPF pathogenesis.

In the present study, we performed computational deconvolution with bulk lung homogenate RNA-seq data from 1,026 subjects in the Lung Tissue Research Consortium (LTRC). By leveraging the large-scale omics data, we tested the hypothesis that there are specific cell types whose abundance and cell type-specific gene expression are associated with disease severity in COPD and IPF subjects.

## METHODS

### Study participants

Research subjects undergoing clinically indicated thoracic surgery were recruited to participate in the LTRC, as previously described [12]. The participating centers’ Institutional Review Boards approved the study, and all subjects provided written informed consent.

COPD subjects included in this analysis had forced expiratory volume in one second (FEV_1_) to forced vital capacity (FVC) ratio <0.70 and FEV_1_ % predicted <80%. Spirometric severity was characterized by Global Initiative for Chronic Obstructive Lung Disease spirometry grades 2-4. COPD subjects had either pathological emphysema and no alternative pathological diagnosis (interstitial lung disease (ILD), idiopathic interstitial pneumonias (IIPs), sarcoidosis, constrictive bronchiolitis, cellular hypersensitivity pneumonitis, diffuse alveolar damage, or eosinophilic granuloma). Any individual meeting the physiological diagnostic criteria for COPD but with a clinical diagnosis of IPF or sarcoidosis was excluded from the COPD group.

IPF subjects had a clinical diagnosis of IPF based on the site’s multidisciplinary diagnostic process of all available data instituted at each participating institution. Control subjects had normal spirometry with no pathologic diagnosis of ILD/IIPs, sarcoidosis, constrictive bronchiolitis, cellular hypersensitivity pneumonitis, diffuse alveolar damage, or eosinophilic granuloma.

### Computational deconvolution

Computational deconvolution was performed using CIBERSORTx (available at https://cibersortx.stanford.edu/) [13]. The docker image obtained from CIBERSORTx website was used with the Podman container image management engine on the Channing Division of Network Medicine GPU computing cluster. This provided computational efficiency beyond what was available through the CIBERSORTx web interface.

We used LTRC TOPMed Harmonized phenotype data set dated November 30, 2022 and freeze 1 LTRC gene expression data set. Data are available on the NCBI database of Genotypes and Phenotypes (dbGaP), accession phs001662 (LTRC). LTRC RNA-seq data from TOPMed (https://topmed.nhlbi.nih.gov) are available through dbGaP. For the count matrix generation, isoform-level expression quantification was generated with Salmon (v1.3.0) pseudoalignment to GENCODE release 37 transcriptome and summarized to gene-level counts using tximeta (v1.8.5). For salmon alignment, seq_bias_correct and gc_bias_correct were set to TRUE. Deconvolution was performed on the entire LTRC dataset that passed RNAseq QC (n=1,555), irrespective of whether the subject was included in our final analysis. Batch effects (library preparation batch) were removed using Combat_seq in the sva R package, and the matrix was cpm normalized after batch effect removal. Genes that had cpm >1 in at least 20% of the LTRC dataset and had assigned HUGO Gene Nomenclature Committee symbols were used in the deconvolution.

In total, 23,097 genes were included in the deconvolution after the batch effect removal and filtering steps. A custom signature matrix from a reference scRNA-seq was generated using CIBERSORTx. The signature matrix is a specialized expression matrix of cell type-specific “barcode” genes which provides a reference atlas of known cellular signatures for the deconvolution procedure. For this process, the CIBERSORTx algorithm used scRNA-seq data on 31,943 lung cells from 44 ever-smokers: six control, seventeen COPD, and twenty-one IPF subjects)[9,10]. Of the 23,097 genes in the LTRC dataset, 19,655 were also in the scRNA-seq dataset (42,406 features across 161,067 cells in the qc’ed dataset); these genes were used to train the CIBERSORTx algorithm. The CIBERSORTx signature matrix we generated is attached as a supplementary file. Thirty-eight discrete cell varieties were queried in the deconvolution; cells were labeled as per Adams et al (Supplemental Table E1) [9]. We chose to use this dataset for two main reasons: 1.) the dataset included a wide range of control, COPD, and IPF subjects. 2.) the dataset included disease-specific cell types such as aberrant basaloid cells. Moreover, the cell annotations for the scRNA-seq were shown to be consistent with automated annotation drawn from multiple cell type definition databases such as the Human Primary Cell Atlas and Blue ENCODE databases, as previously reported [9].

For the imputation of cell fraction, we used CIBERSORTx in fraction mode with single-cell mode set to TRUE and rmbatchSmode set to FALSE; i.e., batch correction and quantile normalization by the CIBERSORTx algorithm were disabled. Proportions were calculated for each sample with all the cell types proportions added up to 1. For deriving abundance scores for cell types, the computation was performed on the CIBERSORTx web interface as this specific function is disabled by the algorithm provided by the authors inside the docker image.

CIBERSORTx estimates the relative fraction of each cell type included in the signature matrix, such that the sum of all fractions is equal to 1 for a given bulk RNA-seq sample. Therefore, the number of cell types included in the signature matrix may impact the relative fraction of each cell type. To overcome this issue, we used CIBERSORTx absolute mode where the absolute abundance score was estimated by the median expression level of all genes in the signature matrix (matrix generated using the reference scRNA-seq matrix) divided by the median expression level of all genes in the sample mixture (LTRC gene expression) [14,15]. This approach allows relative abundance comparisons across samples and cell types. Cell type-specific gene expression matrices were generated using CIBERSORTx high-resolution mode using the docker image and used in the subsequent analyses.

### Cell type-specific differential gene expression analysis

We performed differential gene expression analysis in cell type-specific gene expression matrices to find out which genes, even after removing the cellular abundance effects, were differentially expressed between case and controls. Using cell type-specific gene expression matrices (gene-by-sample matrices for each cell type) generated from CIBERSORTx, we performed differential gene expression analysis using limma [16]. Cell type-specific differential gene expression was log_2_-transformed, and we included only the genes with varying levels in our analysis (a built-in function of CIBERSORTx). We tested the association between cell type-specific gene expression and disease severity separately in the COPD and IPF groups. In COPD subjects, disease severity was measured by lung function tests including forced expiratory volume in 1 second (FEV_1_) and diffusing capacity of the lungs for carbon monoxide as a percent predicted (D_L_CO %). In IPF subjects, disease severity was measured by forced vital capacity (FVC) and D_L_CO %. Linear models were adjusted for age, sex, height, ever smoking, and lifetime smoking intensity (in pack-years). Multiple testing correction was performed by the Benjamini-Hochberg procedure. Significance was determined at a false discovery rate (FDR) of 5%.

### Functional enrichment analysis

We performed functional enrichment analysis using the STRING database version 12.0 (https://string-db.org) [17]. The reason for using STRING was to use a complementary method based on publicly available dataset to explore the functional consequences of differentially expressed genes. Alongside the protein-protein interaction, we also report gene set enrichment results performed using cell type-specific gene expression data which is part of the STRING interactive online platform.

Using the STRING interactive online platform, we queried active interaction sources and obtained confidence value in functional protein-protein interactions for protein network construction. We excluded any protein-protein interaction source that was based on text mining to reduce false positive signals. Active interaction sources include experiments, databases, co-expression, neighborhood, gene fusion, and co-occurrence. The list of genes used in the functional enrichment analysis are included in the Supplemental Table E2 and E3.

## RESULTS

### Subjects

465 subjects met the case criteria for COPD, 213 subjects met the case criteria for IPF, and 348 subjects met the control criteria. Demographic and clinical characteristics of the 1,026 subjects included in our analysis are shown in Table 1. Notably, IPF subjects were predominantly male (70%). The cohort included 90% of self-identified white subjects. COPD subjects were predominantly smokers (95.2% have ever smoked) and IPF and control subjects were 65.3% and 67.8% ever smokers, respectively.

**Table 1:** LTRC subject demographics and lung function tests.

|  | Control | COPD | IPF |
| --- | --- | --- | --- |
| n | 348 | 465 | 213 |
| Age (mean (SD)) | 61.51 (12.53) | 63.35 (9.18) | 63.55 (8.37) |
| Sex = Female sex (%) | 211 (60.6) | 210 (45.2) | 64 (30.0) |
| Race (%) |  |  |  |
| White | 314 (90.2) | 423 (91.0) | 191 (89.7) |
| Asian | 0 ( 0.0) | 0 ( 0.0) | 4 ( 1.9) |
| Black | 22 ( 6.3) | 29 ( 6.2) | 8 ( 3.8) |
| Hispanic | 10 ( 2.9) | 9 ( 1.9) | 4 ( 1.9) |
| Other race | 2 ( 0.6) | 4 ( 0.9) | 6 ( 2.8) |
| BMI (mean (SD)) | 28.95 (5.97) | 26.30 (5.22) | 29.80 (5.44) |
| Ever Smoking (%) | 215 (67.8) | 415 (95.2) | 128 (65.3) |
| Pack years of smoking (mean (SD)) | 20.12 (27.32) | 47.16 (31.73) | 18.84 (24.18) |
| FEV1/FVC (mean (SD)) | 0.77 (0.06) | 0.45 (0.15) | 0.83 (0.07) |
| FEV1 pp (mean (SD)) | 95.87 (12.61) | 41.72 (20.27) | 65.91 (19.06) |
| FVC pp (mean (SD)) | 96.09 (12.69) | 68.36 (18.50) | 60.28 (17.75) |
| DLCO % (mean (SD)) | 73.22 (15.44) | 42.92 (18.87) | 38.04 (19.59) |

Abbreviations: COPD, chronic obstructive pulmonary disease; IPF, idiopathic pulmonary fibrosis; SD, standard deviation; BMI, body mass index; FEV<sub>1</sub>, forced expiratory volume in 1s; FVC, forced vital capacity; DLCO, diffusing capacity of the lungs for carbon monoxide as a percent predicted.

### Cellular composition differences among COPD, IPF, and controls

Of the thirty-eight cell types queried in the deconvolution, twenty-seven cell types were detected in at least 10% of samples. Of these, there were nineteen cell types whose median proportion was greater than 1% in any one of the groups, as shown in Figure 1.

**Figure 1.**
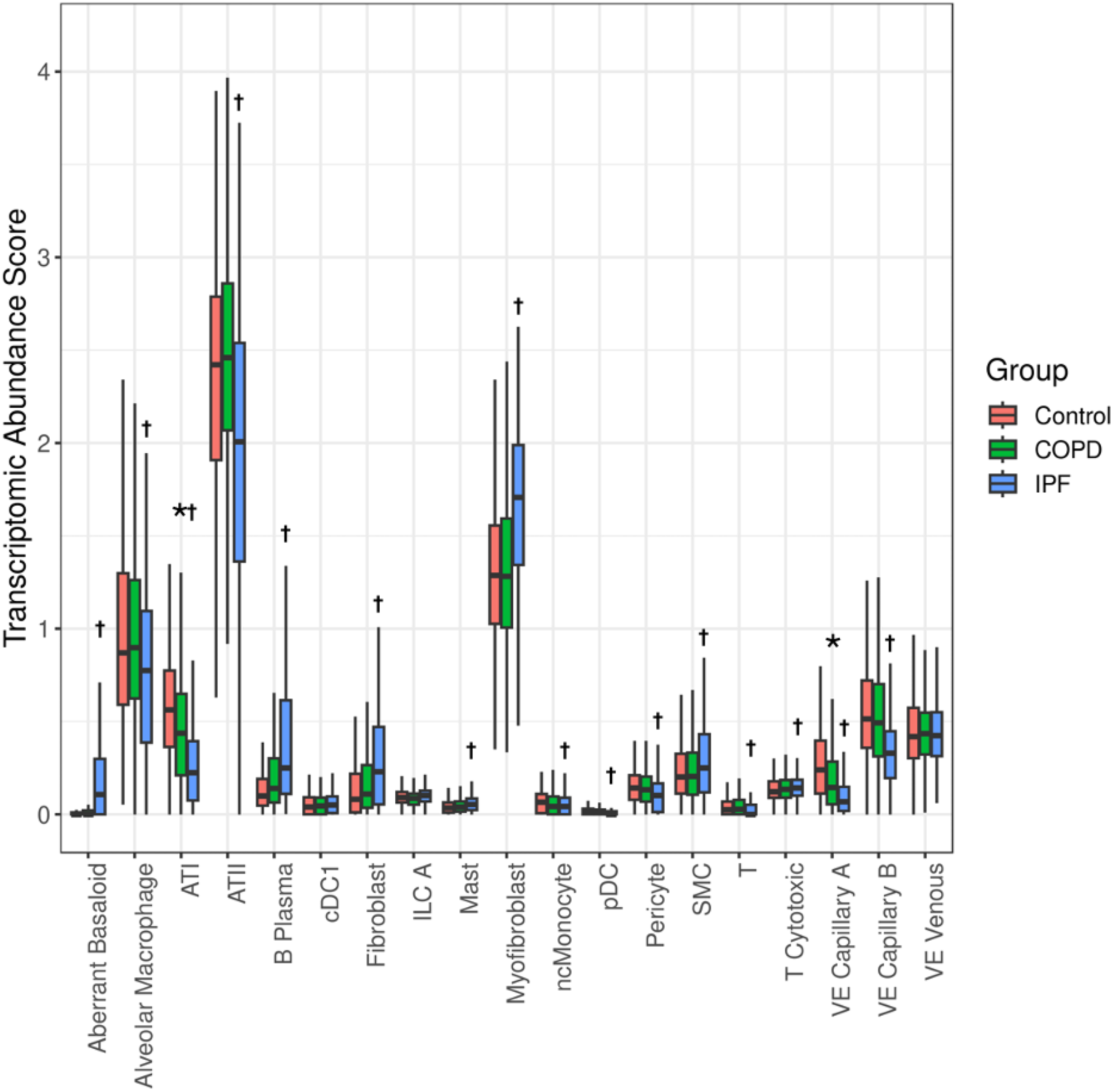
Boxplots showing cell type abundance score for each cell type split by disease status. Results are shown only for cell types detected in at least 10% of samples and whoe median proportion was greater than 1%. Statistical comparison was tested using linear regression adjusting for age, sex, ever smoking and total pack-year. * and † denote significant difference between COPD vs control and IPF vs control, respectively. Abbreviations: COPD, chronic obstructive pulmonary disease; IPF, idiopathic pulmonary fibrosis; ATI, alveolar epithelial type 1 cells; ATII, alveolar epithelial type 2 cells; cDC, classical dendritic cells; ILC, innate lymphoid cells; pDC, plasmacytoid dendritic cells; ncMonocyte, non-classical monocytesSMC; smooth muscle cells; VE Capillary A, vascular endothelial - aerocyte capillary; VE Capillary B, vascular endothelial - general capillary; VE Venous, venous vascular endothelial.

We compared the cell abundance score between COPD, IPF, and control subjects, adjusting for age, sex, height, ever smoking, and smoking pack-years. Figure 1 summarizes cell types whose abundance scores were significantly different (FDR<0.05) between COPD and control subjects and between IPF and control subjects, respectively. VE Capillary A and ATI were lower in COPD tissue compared to controls. Nine cell types were decreased and eleven were increased in IPF compared to controls.

### Associations between cell-type abundance and disease severity in COPD and IPF lungs

Next, we identified cell types whose abundance scores in COPD and IPF lungs were associated with disease severity measured by FEV_1_ (COPD), FVC (IPF), and D_L_CO (COPD and IPF). In COPD subjects, there were two and six cell types that were significantly associated with FEV_1_ and D_L_CO, respectively (Table 2). In IPF subjects, there were eleven and nine cell types that were significantly associated with FVC and D_L_CO, respectively (Table 2). Decreases in the abundances of type A capillary vascular endothelial cells and non-classical monocytes were associated with worse disease severity in both COPD and IPF subjects. In IPF, aberrant basaloid cells showed the strongest association with both FVC and D_L_CO. In fact, we performed additional analysis testing the association between cell abundance score and GAP index [18], a mortality predictive score based on gender (G), age (A), and physiological measures (P; FVC, and D_L_CO) in IPF, and found that aberrant basaloid cells had one of the strongest associations with the index (Supplemental Table E4).

**Table 2:** Cell-type transcriptome abundance score associated with disease severity in COPD and IPF.

| Disease | Outcome | Cell type | Beta | 95% CI | Adjusted p value |
| --- | --- | --- | --- | --- | --- |
| COPD | FEV<sub>1</sub> | VE Capillary A | 0.11 | 0.06,0.16 | 0.001 |
| COPD | FEV<sub>1</sub> | ncMonocyte | 0.09 | 0.03,0.14 | 0.02 |
| COPD | D<sub>L</sub>CO | ATI | 4.79 | 2.99,6.59 | <0.001 |
| COPD | D<sub>L</sub>CO | Macrophage | -4.75 | -7.02,-2.49 | <0.001 |
| COPD | D<sub>L</sub>CO | ncMonocyte | 3.71 | 1.94,5.49 | <0.001 |
| COPD | D<sub>L</sub>CO | VE Capillary A | 3.69 | 1.95,5.44 | <0.001 |
| COPD | D<sub>L</sub>CO | cMonocyte | -3.16 | -5.47,-0.86 | 0.031 |
| COPD | D<sub>L</sub>CO | ILC A | 2.72 | 0.93,4.51 | 0.016 |
| IPF | FVC | ncMonocyte | 0.31 | 0.21,0.4 | <0.001 |
| IPF | FVC | Aberrant Basaloid | -0.24 | -0.34,-0.14 | <0.001 |
| IPF | FVC | Macrophage | -0.20 | -0.31,-0.1 | 0.001 |
| IPF | FVC | cMonocyte | -0.19 | -0.29,-0.09 | 0.001 |
| IPF | FVC | T Cytotoxic | 0.19 | 0.09,0.29 | 0.002 |
| IPF | FVC | ATII | 0.16 | 0.06,0.27 | 0.007 |
| IPF | FVC | VE Venous | -0.16 | -0.27,-0.06 | 0.012 |
| IPF | FVC | Alveolar Macrophage | 0.15 | 0.05,0.25 | 0.016 |
| IPF | FVC | VE Capillary A | 0.15 | 0.05,0.26 | 0.016 |
| IPF | FVC | T | -0.14 | -0.24,-0.03 | 0.029 |
| IPF | FVC | pDC | -0.12 | -0.23,-0.02 | 0.046 |
| IPF | D<sub>L</sub>CO | Aberrant Basaloid | -6.42 | -9.25,-3.58 | <0.001 |
| IPF | D<sub>L</sub>CO | Alveolar Macrophage | 6.08 | 3.19,8.97 | 0.001 |
| IPF | D<sub>L</sub>CO | ATII | 5.65 | 2.8,8.51 | 0.001 |
| IPF | D<sub>L</sub>CO | T Cytotoxic | 5.07 | 2.04,8.1 | 0.006 |
| IPF | D<sub>L</sub>CO | VE Capillary A | 5.06 | 2.1,8.03 | 0.006 |
| IPF | D<sub>L</sub>CO | ncMonocyte | 4.79 | 1.89,7.69 | 0.006 |
| IPF | D<sub>L</sub>CO | Macrophage | -4.64 | -7.64,-1.65 | 0.01 |
| IPF | D<sub>L</sub>CO | cMonocyte | -4.34 | -7.53,-1.15 | 0.023 |
| IPF | D<sub>L</sub>CO | T Regulatory | 3.92 | 1.06,6.77 | 0.023 |

Statistical comparison was tested using linear regression adjusting for age, sex, height, ever smoking and total pack-year. Beta was estimated using absolute value of outcome measures and are estimated per one standard deviation change in CIBERSORTx absolute abundance score. Pre-bronchodilator FEV<sub>1</sub> and DLCO percent predicted were used. Abbreviations: COPD, chronic
obstructive pulmonary disease; IPF, idiopathic pulmonary fibrosis; CI, confidence interval; FEV<sub>1</sub>, forced expiratory volume in 1s; FVC, forced vital capacity; DLCO, diffusing capacity of the lungs for carbon monoxide as a percent predicted; VE Capillary A, vascular endothelial -aerocyte capillary; ncMonocyte, non-classical monocytes; cMonocyte, classical monocytes; ATI, alveolar epithelial type 1 cells; ILC A, type A innate lymphoid cells; ATII, alveolar epithelial type 2 cells; VE Venous, vascular endothelial venous cells; pDC, plasmacytoid dendritic cells.

### Associations between cell type-specific gene expression and disease severity in COPD and IPF lungs

We estimated cell type-specific gene expression for cell types whose median proportion was greater than 1%: ATII, Alveolar Macrophage, SMC, Fibroblast, ATI, Myofibroblast, VE Capillary B, B Plasma, VE Capillary A, ILC A, VE Venous, Pericyte, and T Cytotoxic. Table 3 summarizes the number of differentially expressed genes in COPD and IPF. Overall, there were more differentially expressed genes (FDR<0.05) in IPF lungs than in COPD lungs. ATII cells and alveolar macrophages were two cell types with the greatest number of genes with cell type-specific differential gene expression associated with disease severity in both diseases. Aberrant basaloid cells, despite being estimated to represent only 1.3 % (IQR: 0-3.5 %) of cell proportion in IPF subjects, had the second largest number of cell type-specific genes whose expression was positively associated with IPF severity.

**Table 3:** Cell type-specific differential gene expression in COPD and IPF lungs compared to control lungs.

| Cell type | Total genes in analysis | Number of upregulated genes in COPD | Number of downregulated genes in COPD | Number of upregulated genes in IPF | Number of downregulated genes in IPF |
| --- | --- | --- | --- | --- | --- |
| ATII | 10964 | 272 | 2088 | 3886 | 2967 |
| Alveolar Macrophage | 5614 | 228 | 772 | 1847 | 1275 |
| SMC | 4265 | 40 | 183 | 1922 | 604 |
| Fibroblast | 3500 | 89 | 254 | 1268 | 658 |
| ATI | 3362 | 252 | 108 | 819 | 578 |
| Myofibroblast | 3099 | 120 | 399 | 1225 | 612 |
| VE Capillary B | 2446 | 54 | 338 | 412 | 943 |
| B Plasma | 2325 | 116 | 50 | 1039 | 460 |
| VE Capillary A | 2063 | 73 | 284 | 404 | 669 |
| ILC A | 2049 | 8 | 1 | 332 | 240 |
| VE Venous | 1310 | 33 | 114 | 230 | 374 |
| Pericyte | 1210 | 26 | 56 | 326 | 275 |
| T Cytotoxic | 1139 | 7 | 2 | 192 | 81 |

Associations were tested using limma [16] on variable genes only. Significant association were adjusted to FDR 5%. Abbreviations: COPD, chronic obstructive pulmonary disease; IPF, idiopathic pulmonary fibrosis; ATII, alveolar epithelial type 2 cells; SMC, smooth muscle cells; ATI, alveolar epithelial type 1 cells; VE Capillary B, vascular endothelial - general capillary; VE Capillary A, vascular endothelial - aerocyte capillary; ILC A, type A innate lymphoid cells; VE Venous, vascular endothelial venous cells.

Next, we tested the association between cell type-specific gene expression and disease severity in COPD and IPF subjects. We included all cell types whose median proportion was greater than 1% in each disease group. In COPD subjects, cell types tested were Alveolar Macrophage, ATI, ATII, B Plasma, Fibroblast, ILC A, Myofibroblast, Pericyte, SMC, T Cytotoxic, VE Capillary A, VE Capillary B, and VE Venous. In IPF subjects, cell types tested included Aberrant Basaloid, Alveolar Macrophage, ATI, ATII, B Plasma, Fibroblast, ILC A, Myofibroblast, Pericyte, SMC, T Cytotoxic, VE Capillary B, and VE Venous. Figure 2 (and Figure E2) shows the number of genes with cell type-specific expression associated with lung function measures in COPD and IPF subjects. We also provide a list of all cell type-specific gene expression associations with disease severity in IPF and COPD (Supplemental Table E5 and 6). Figure 2 also shows the number of genes with cell that overlap between the two different measures of disease severity.

**Figure 2.**
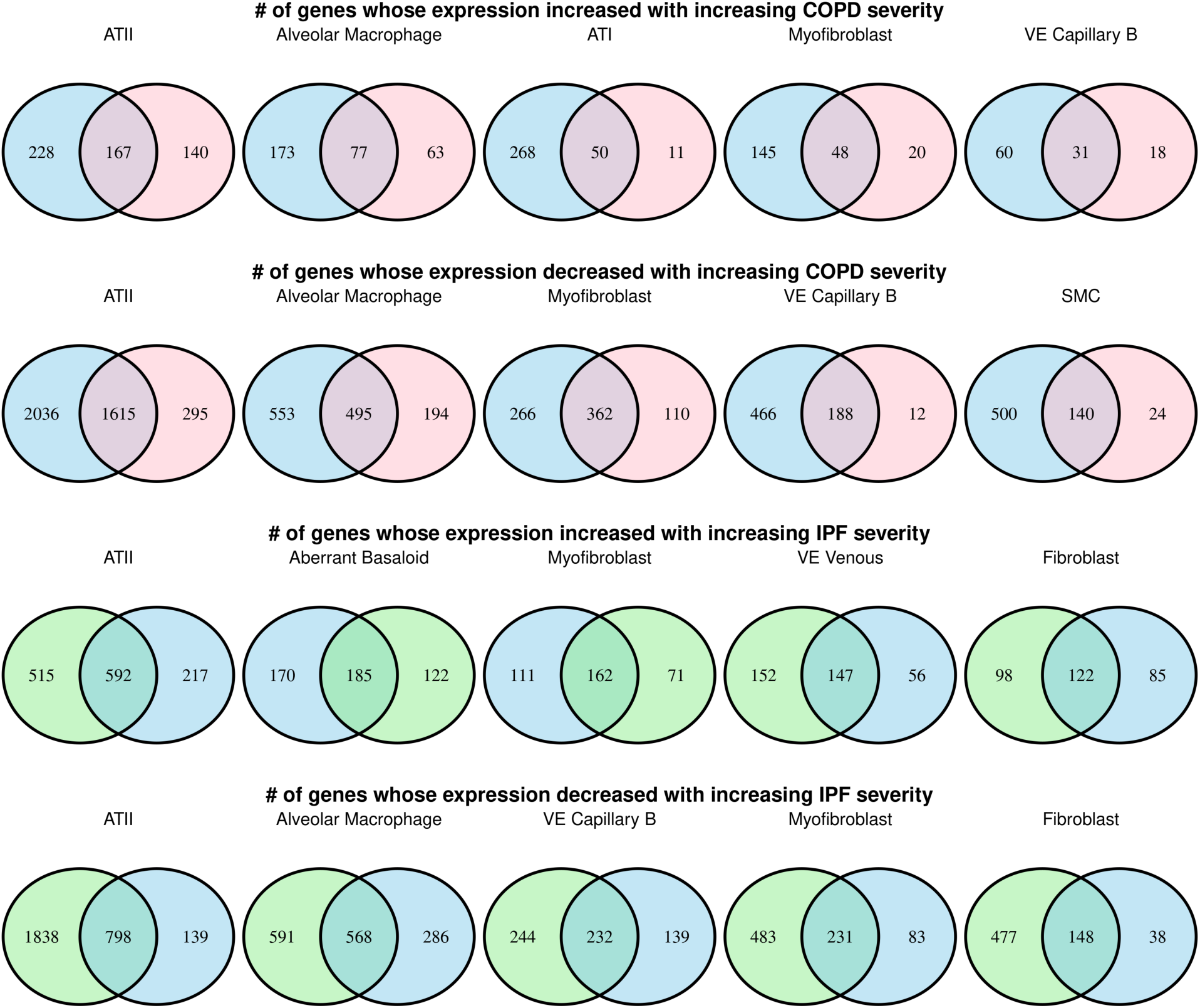
Venn diagrams showing the five cell types with the most cell type-specific gene expression levels associated with disease severity in COPD and IPF lungs. Genes associated with DLCO, FEV1, and FVC are colored blue, pink, and green, respectively. Cell-types with a higher number of gene expressions associated with disease severity are ordered left to right.

Supplemental Tables E7 and E8 summarize the number of significant cell type-specific gene expressions associated with disease severity in COPD and IPF, respectively. Of note, besides the ATII cells, which were the most abundant cell types in the samples estimated using RNA-seq deconvolution, alveolar macrophages in COPD and aberrant basaloid cells had the highest number of genes associated with disease severity in both COPD and IPF. Hence, we chose these two cell types to perform functional enrichment analyses and highlight their upregulated function.

### Functional enrichment analysis of genes associated with COPD severity in alveolar macrophages

We performed functional enrichment analysis using the list of genes whose expression levels in alveolar macrophages were positively associated with COPD severity as measured by FEV_1_ and D_L_CO. We queried all matched proteins encoded by the 77 genes identified in this cell type-specific differential gene expression analysis. In the protein-protein interaction (PPI) network analysis in the STRING database, we found significant functional enrichment with 144 edges (expected number of edges 60; PPI enrichment p-value <1×10^-16^). Figure 3 shows the PPI network for proteins encoded by the alveolar macrophage gene expression that is positively associated with COPD severity. The result of the functional enrichment analysis is included in the online Supplement Table E8. The most significantly enriched term was from the Reactome database for Eukaryotic Translation Elongation (Reactome term HSA-156842: FDR = 1.25 × 10^-11^).

**Figure 3.**
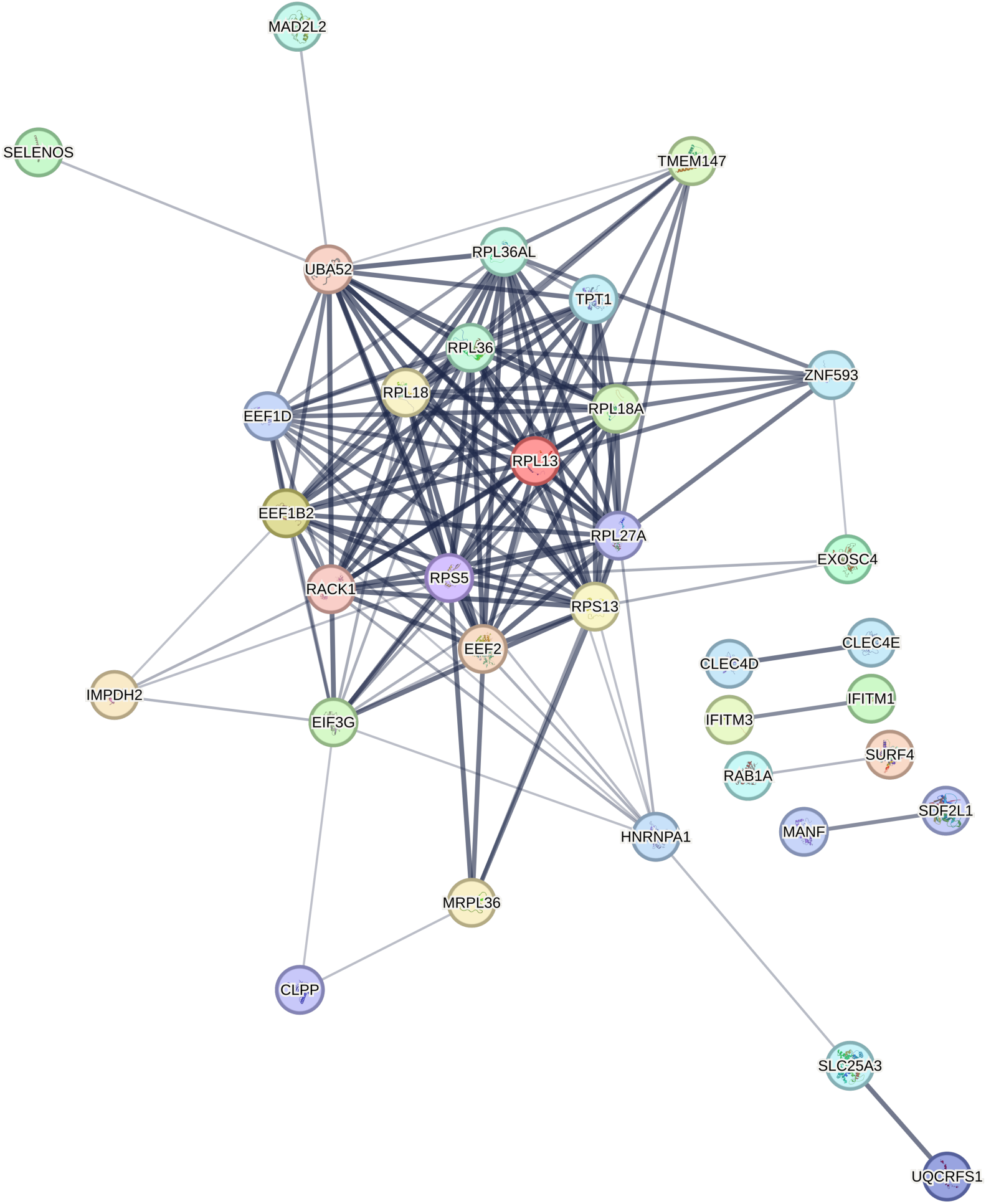
Protein-protein interaction network for the proteins encoded by genes in alveolar macrophages positively associated with COPD severity. Edges represent protein-protein associations based on association confidence score calculated using STRING database (version 12.0). The edge line thickness indicates the strength of data support. Disconnected nodes in the network were hidden for illustrative purpose.

### Functional enrichment analysis of genes associated with IPF severity in aberrant basaloid cells

We performed functional enrichment analysis using the list of genes whose expression levels in aberrant basaloid cells were positively associated with IPF severity as measured by FVC and D_L_CO. We queried all matched proteins encoded by the 185 genes identified in the cell type-specific differential gene expression analysis. We found significant functional enrichment with 123 edges (expected number of edges 53; PPI enrichment p-value = 2.22×10^-16^). Figure 4 shows the PPI network for proteins encoded by the aberrant basaloid genes positively associated with IPF severity. The result of the functional enrichment analysis is included in the Online Supplement Table E9. Formation of the cornified envelope (STRING Cluster ID CL34114; FDR = 5.85 × 10^-14^) was indicated as the top most significant functional enrichment term.

**Figure 4.**
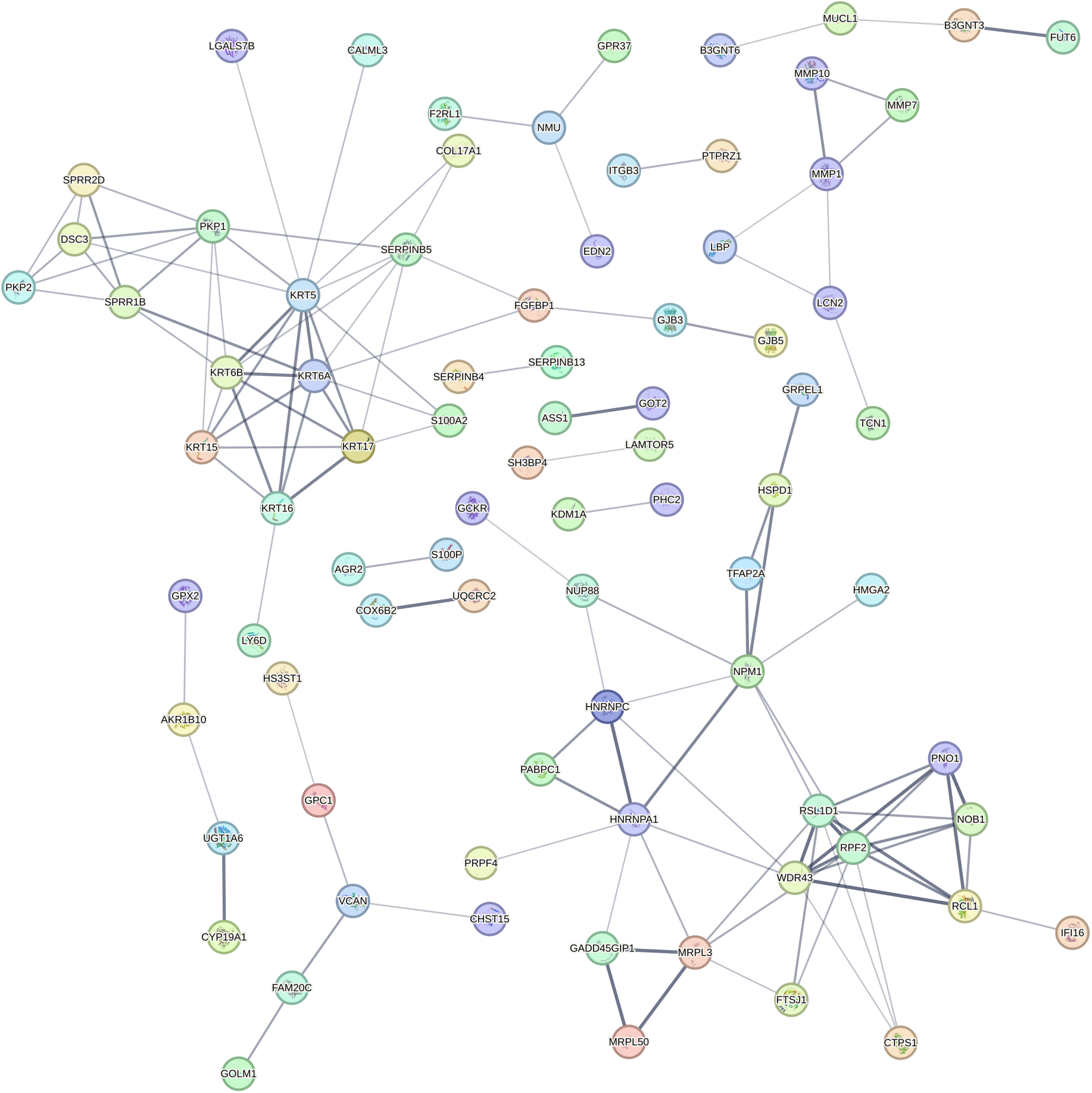
Protein-protein interaction network for proteins encoded by genes in aberrant basaloid cells that were positively associated with IPF severity. Edges represent protein-protein associations based on association confidence score calculated using STRING database (version 12.0). The edge line thickness indicates the strength of data support. Disconnected nodes in the network were hidden for illustrative purpose.

## DISCUSSION

We report the results of a computational tissue profiling analysis of bulk lung RNA-seq data from 1,026 subjects in the LTRC. We report the cellular composition and cell type-specific gene expression in lung tissue associated with disease severity in COPD and IPF subjects, extending the single-cell experiment discoveries from a modest sample size (<100 subjects) to a large population cohort (>1000 subjects). We trained a well-established and widely implemented computational RNA-seq deconvolution algorithm, CIBERSORTx [13,19,20], using publicly available scRNA-seq data from control, COPD, and IPF subjects [9].

We found that IPF lung tissues showed the most divergence from control lungs in cellular composition, with eighteen cell types whose abundance score was different from the controls, adjusting for covariates. Our results showed in a large IPF sample the association of aberrant basaloid cells and their expression with IPF and IPF severity; the association with IPF severity has not been previously reported. We also found that abundances of eight cell types— ncMonocyte, Aberrant Basaloid, Macrophage, cMonocyte, T Cytotoxic, ATII, Alveolar Macrophage, and VE Capillary A—were associated with disease severity in the IPF subjects. Structural cells such ATII, aberrant basaloid cells, myofibroblasts, and fibroblasts were among the cell types with the most number of genes associated with IPF severity. Notably, we found that aberrant basaloid cells were enriched in IPF lungs, and that the abundance of this disease-enriched cell type increased as the disease severity increased. It is notable that aberrant basaloid proportions remained below 1% in COPD.

In aberrant basaloid cells, expression levels of matrix metallopeptidase 7 (*MMP7*), growth differentiation factor 15 (*GDF15*), and eph receptor B2 (*EPHB2*), were negatively associated with FVC or D_L_CO. In other words, the expression of these genes increased in more severe disease. These genes and the protein they encode have been implicated in the pathogenesis of IPF [21–24]. Our data supports the notion that GDF15 may be circulating biomarker reflective of aberrant basaloid cells in the airway epithelium [23]. We also found that *EPHB2* level in myofibroblasts was positively associated with IPF severity, extending the previous scRNA-seq finding that demonstrated increase level of *EPHB2* in IPF subjects compared to controls [9].

The functional enrichment analysis showed that the formation of the cornified envelope and keratinization were functionally enriched in aberrant basaloid cells with increasing severity of IPF. The cornified cell envelope is a highly insoluble and extremely tough structure that forms under the epithelium to help the epithelium defend against reactive oxygen species [25]. This may result from and/or be a contributing factor to the tissue fibrosis in IPF; however, alteration in this cellular function has not been implicated in IPF previously. Therefore, this result will require further validation at the protein level. In addition to these functions, the protein interaction network analysis also highlighted the increased expression of matrix metalloproteases such as *MMP7*, *MMP10*, and *MMP1*, along with their functionally associated genes such as lipopolysaccharide binding protein (*LBP*), lipocalin 2 (*LCN2*), and transcobalamin1 (*TCN1*), in aberrant basaloid cells with increased disease severity. These results suggest that increased abundance of aberrant basaloid cells and their gene expression of cellular processes involved in aberrant barrier formation and extracellular matrix modification is associated with IPF severity.

We also showed that cellular composition is different between COPD and controls and that there were several cell types whose abundance was associated with COPD severity. There was a significant decrease in alveolar type 1 cells and capillary type A vascular endothelial cells in COPD lungs compared to controls. Capillary type A vascular endothelial cells were also negatively associated with increasing disease severity as measured by FEV_1_ and D_L_CO. This observation provides additional evidence linking endothelial injury to COPD and extends earlier findings that identified injury to pulmonary vessels in lung tissue from COPD patients [26].

Beyond the pulmonary vasculature, the abundance of macrophage, ncMonocyte, and cMonocyte were associated with D_L_CO, but only ncMonocytes abundance was significantly associated with FEV_1_.

Monocytes and macrophages play an important role in pulmonary host defenses through their phagocytic activities and regulation of innate and adaptive immunity. The circulating monocyte pool and macrophages in tissue are composed of multiple subsets, each with a specialized function. Animal models and human *ex vivo* experiments have demonstrated the dysregulated functions of macrophage populations in COPD lungs [27]. Extensive molecular characterizations of immune cells in COPD, particularly the lung macrophage populations, have been conducted using flow cytometry and other low-throughput molecular techniques [28–30]. However, due to the practicality of needing fresh samples and the experimental cost, tissue and immune profiling studies have been limited in terms of sample sizes (typically <100 subjects) and the small number of molecular targets. Recently, scRNA-seq studies with more molecular targets have been conducted and highlighted immunological dysregulation of monocytes and macrophages in COPD [9,10,31,32]. However, the number of COPD donors was small in these studies, and there was limited information on the disease phenotypes, which limited the ability to test for associations with disease severity, clinical outcomes, and pathological changes. Our computational tissue profiling in a large-scale cohort builds on this important body of work and extends the findings from scRNA-seq to an epidemiological cohort.

Given the important role alveolar macrophages play in COPD pathogenesis, we focused on this cell type for functional enrichment analysis, which highlighted that increased disease severity was associated with increased mRNA encoding for proteins involved in translation and energy metabolism. This finding agrees with previous studies that macrophage metabolic function is associated with COPD and supports the notion that metabolomic reprogramming of lung macrophages is important in the pathogenesis of COPD [33,34]. We provide the list of cell type-specific genes associated with COPD and IPF severity (Supplemental Table E5 and E6) for the community to explore using the cell type-specific functional enrichment using tools such as STRING database (https://string-db.org/) for other cell types.

There are some limitations of our study. First, RNA-seq based deconvolution methods are more suited for analysis of highly abundant cell types (cell types with frequency >1%) [13,19]. It is also influenced by the size of the cell type-specific transcriptome. This makes rare cell types with small transcriptomes challenging to study using the deconvolution approach. To overcome this issue, future studies may combine RNA-seq deconvolution with results based on other omics (e.g., DNA methylation-based deconvolution). Also, careful enrichment of a cell type by FACS sorting may be required to study rarer cell populations. Second, bulk tissue analysis is limited in spatial resolution. This limits the understanding of the spatial distribution and interaction of cells in the diseased lungs. Nevertheless, our study informs which cell types may be the better candidates to be the focus of future spatial transcriptomic investigations. Finally, the study was limited to a population of predominantly white subjects with access to U.S. academic medical centers. This may limit the generalizability and calls for future efforts to include subjects from multi-ethnic and multi-national backgrounds.

## CONCLUSION

In conclusion, we present here the cellular composition changes and cell type-specific gene expression associated with disease severity in COPD and IPF lungs. We document the cell types whose estimated abundance is associated with the severity of disease in COPD or IPF. We highlight two cell types—alveolar macrophages in COPD and aberrant basaloid cells in IPF— whose cell type-specific gene expressions were associated with clinical measures of disease severity. We also highlight the cell type-specific functional enrichment pointing to the altered cellular functions associated with disease severity. Using computational deconvolution, this study extends single-cell experimental discoveries from a modest sample size to a large population cohort and contributes to our understanding of tissue heterogeneity in COPD and IPF pathobiology. This knowledge offers insight into the alterations within lung tissue in advanced illness, providing a better understanding of the underlying pathological processes that drive disease progression.

## List of Abbreviations

ATI: alveolar type 1 pneumocytes
ATII: alveolar type 2 pneumocytes
COPD: chronic obstructive pulmonary disease
DLCO: diffusing capacity for carbon monoxide
FACS: fluorescence-activated cell sorting
FDR: false discovery rate
FEV1: forced expiratory volume in one second
FVC: forced vital capacity
IIPs: idiopathic interstitial pneumonias
ILC A: type A innate lymphoid cells
ILD: interstitial lung disease
IPF: idiopathic pulmonary fibrosis
IQR: interquartile range
LTRC: Lung Tissue Research Consortium
PPI: protein-protein interaction
RNA-seq: RNA sequencing
SMC: smooth muscle cells
SMC: smooth muscle cells
TGF-β: transforming growth factor beta
VE Capillary A: vascular endothelial - aerocyte capillary
VE Capillary B: vascular endothelial - general capillary
VE Venous: vascular endothelial venous cells
cMonocyte: classical monocytes
ncMonocyte: non-classical monocytes
pDC: plasmacytoid dendritic cells, and
scRNA-seq: single-cell RNA sequencing

## DECLARATIONS

### Ethics approval and consent to participate

The participating centers’ Institutional Review Boards approved the study, and all subjects provided written informed consent.

### Consent for publication

Not applicable.

### Availability of data and materials

Data are available on the NCBI database of Genotypes and Phenotypes (dbGaP), accession phs001662 (LTRC). LTRC RNA-seq data from TOPMed (https://topmed.nhlbi.nih.gov) are available through dbGaP. The analysis results and code can be obtained by contacting the corresponding author with a reasonable request.

## Author contributions

Concept and design: MHR, CPH, and JDM; data collection: JHY, FS, LB, AL, GC, KB, RW, FM, KF, MHC, PJC, DLD, EKS, CPH, and JDM; statistical support: MHR, KJK, MG, CPH, JDM; data analysis: MHR, JHY, KJK, MG, AG, and JDM; manuscript writing - draft: MHR, CPH, and JDM; manuscript writing - edit: all authors; funding: PJC, EKS, and CPH. All authors read and approved the final manuscript.

## Acknowledgements

NHLBI TOPMed: Lung Tissue Research Consortium Molecular data from the Trans-Omics in Precision Medicine (TOPMed) program was supported by the National Heart, Lung, and Blood Institute (NHLBI). RNASeq for “NHLBI TOPMed: Lung Tissue Research Consortium” (phs001662) was performed at the Northwest Genomics Center (HHSN268201600032I). Core support including centralized genomic read mapping and genotype calling, along with variant quality metrics and filtering were provided by the TOPMed Informatics Research Center (3R01HL-117626-02S1; contract HHSN268201800002I). Core support including phenotype harmonization, data management, sample-identity QC, and general program coordination were provided by the TOPMed Data Coordinating Center (R01HL-120393; U01HL-120393; contract HHSN268201800001I). We gratefully acknowledge the studies and participants who provided biological samples and data for TOPMed.

## REFERENCES

[1] Agustí A, Hogg JC. Update on the pathogenesis of chronic obstructive pulmonary disease. New England Journal of Medicine 2019;381:1248–56. 10.1056/nejmra1900475.

[2] Lederer DJ, Martinez FJ. Idiopathic pulmonary fibrosis. New England Journal of Medicine 2018;378:1811–23. 10.1056/nejmra1705751.

[3] Selman M, Martinez FJ, Pardo A. Why does an aging smoker’s lung develop idiopathic pulmonary fibrosis and not chronic obstructive pulmonary disease? American Journal of Respiratory and Critical Care Medicine 2019;199:279–85. 10.1164/rccm.201806-1166pp.

[4] Chen S, Kuhn M, Prettner K, Yu F, Yang T, Bärnighausen T, et al. The global economic burden of chronic obstructive pulmonary disease for 204 countries and territories in 202050: a health-augmented macroeconomic modelling study. The Lancet Global Health 2023;11:e1183–93. 10.1016/s2214-109x(23)00217-6.

[5] Wong AW, Koo J, Ryerson CJ, Sadatsafavi M, Chen W. A systematic review on the economic burden of interstitial lung disease and the cost-effectiveness of current therapies. BMC Pulmonary Medicine 2022;22. 10.1186/s12890-022-01922-2.

[6] Sakornsakolpat P, Prokopenko D, Lamontagne M, Reeve NF, Guyatt AL, Jackson VE, et al. Genetic landscape of chronic obstructive pulmonary disease identifies heterogeneous cell-type and phenotype associations. Nature Genetics 2019;51:494–505. 10.1038/s41588-018-0342-2.

[7] Allen RJ, Stockwell A, Oldham JM, Guillen-Guio B, Schwartz DA, Maher TM, et al. Genome-wide association study across five cohorts identifies five novel loci associated with idiopathic pulmonary fibrosis. Thorax 2022;77:829–33. 10.1136/thoraxjnl-2021-218577.

[8] Allen RJ, Guillen-Guio B, Oldham JM, Ma S-F, Dressen A, Paynton ML, et al. Genome-wide association study of susceptibility to idiopathic pulmonary fibrosis. American Journal of Respiratory and Critical Care Medicine 2020;201:564–74. 10.1164/rccm.201905-1017oc.

[9] Adams TS, Schupp JC, Poli S, Ayaub EA, Neumark N, Ahangari F, et al. Single-cell RNA-seq reveals ectopic and aberrant lung-resident cell populations in idiopathic pulmonary fibrosis. Science Advances 2020;6. 10.1126/sciadv.aba1983.

[10] Sauler M, McDonough JE, Adams TS, Kothapalli N, Barnthaler T, Werder RB, et al. Characterization of the COPD alveolar niche using single-cell RNA sequencing. Nature Communications 2022;13. 10.1038/s41467-022-28062-9.

[11] Villaseñor-Altamirano AB, Jain D, Jeong Y, Menon JA, Kamiya M, Haider H, et al. Activation of CD8^+^ T cells in chronic obstructive pulmonary disease lung. American Journal of Respiratory and Critical Care Medicine 2023;208:1177–95. 10.1164/rccm.202305-0924oc.

[12] Yang IV, Pedersen BS, Rabinovich E, Hennessy CE, Davidson EJ, Murphy E, et al. Relationship of DNA methylation and gene expression in idiopathic pulmonary fibrosis. American Journal of Respiratory and Critical Care Medicine 2014;190:1263–72. 10.1164/rccm.201408-1452oc.

[13] Newman AM, Steen CB, Liu CL, Gentles AJ, Chaudhuri AA, Scherer F, et al. Determining cell type abundance and expression from bulk tissues with digital cytometry. Nature Biotechnology 2019;37:773–82. 10.1038/s41587-019-0114-2.

[14] Newman AM, Liu CL, Green MR, Gentles AJ, Feng W, Xu Y, et al. Robust enumeration of cell subsets from tissue expression profiles. Nature Methods 2015;12:453–7. 10.1038/nmeth.3337.

[15] Chen B, Khodadoust MS, Liu CL, Newman AM, Alizadeh AA. Profiling tumor infiltrating immune cells with CIBERSORT, Springer New York; 2018, p. 243–59. 10.1007/978-1-4939-7493-1_12.

[16] Ritchie ME, Phipson B, Wu D, Hu Y, Law CW, Shi W, et al. limma powers differential expression analyses for RNA-sequencing and microarray studies. Nucleic Acids Research 2015;43:e47–7. 10.1093/nar/gkv007.

[17] Szklarczyk D, Kirsch R, Koutrouli M, Nastou K, Mehryary F, Hachilif R, et al. The STRING database in 2023: proteinprotein association networks and functional enrichment analyses for any sequenced genome of interest. Nucleic Acids Research 2022;51:D638–46. 10.1093/nar/gkac1000.

[18] Ley B, Ryerson CJ, Vittinghoff E, Ryu JH, Tomassetti S, Lee JS, et al. A Multidimensional Index and Staging System for Idiopathic Pulmonary Fibrosis. Annals of Internal Medicine 2012;156:684. 10.7326/0003-4819-156-10-201205150-00004.

[19] Jin H, Liu Z. A benchmark for RNA-seq deconvolution analysis under dynamic testing environments. Genome Biology 2021;22. 10.1186/s13059-021-02290-6.

[20] Im Y, Kim Y. A comprehensive overview of RNA ceconvolution methods and their application. Molecules and Cells 2023;46:99–105. 10.14348/molcells.2023.2178.

[21] Bauer Y, White ES, Bernard S de, Cornelisse P, Leconte I, Morganti A, et al. MMP-7 is a predictive biomarker of disease progression in patients with idiopathic pulmonary fibrosis. ERJ Open Research 2017;3:00074–2016. 10.1183/23120541.00074-2016.

[22] Pardo A, Cabrera S, Maldonado M, Selman M. Role of matrix metalloproteinases in the pathogenesis of idiopathic pulmonary fibrosis. Respiratory Research 2016;17. 10.1186/s12931-016-0343-6.

[23] Zhang Y, Jiang M, Nouraie M, Roth MG, Tabib T, Winters S, et al. GDF15 is an epithelial-derived biomarker of idiopathic pulmonary fibrosis. American Journal of Physiology-Lung Cellular and Molecular Physiology 2019;317:L510–21. 10.1152/ajplung.00062.2019.

[24] Lagares D, Ghassemi-Kakroodi P, Tremblay C, Santos A, Probst CK, Franklin A, et al. ADAM10-mediated ephrin-B2 shedding promotes myofibroblast activation and organ fibrosis. Nature Medicine 2017;23:1405–15. 10.1038/nm.4419.

[25] Schäfer M, Werner S. The cornified envelope: a first line of defense against reactive oxygen species. Journal of Investigative Dermatology 2011;131:1409–11. 10.1038/jid.2011.119.

[26] Polverino F, Celli BR, Owen CA. COPD as an endothelial disorder: endothelial injury linking lesions in the lungs and other organs? (2017 Grover Conference Series). Pulmonary Circulation 2018;8:1–18. 10.1177/2045894018758528.

[27] Kapellos TS, Bassler K, Aschenbrenner AC, Fujii W, Schultze JL. Dysregulated functions of lung macrophage populations in COPD. Journal of Immunology Research 2018;2018:1–19. 10.1155/2018/2349045.

[28] Tesfaigzi Y, Curtis JL, Petrache I, Polverino F, Kheradmand F, Adcock IM, et al. Does chronic obstructive pulmonary disease originate from different cell types? American Journal of Respiratory Cell and Molecular Biology 2023;69:500–7. 10.1165/rcmb.2023-0175ps.

[29] Freeman CM, Curtis JL. Lung dendritic cells: shaping immune responses throughout chronic obstructive pulmonary disease progression. American Journal of Respiratory Cell and Molecular Biology 2017;56:152–9. 10.1165/rcmb.2016-0272tr.

[30] Dewhurst JA, Lea S, Hardaker E, Dungwa JV, Ravi AK, Singh D. Characterisation of lung macrophage subpopulations in COPD patients and controls. Scientific Reports 2017;7. 10.1038/s41598-017-07101-2.

[31] Morrow JD, Chase RP, Parker MM, Glass K, Seo M, Divo M, et al. RNA-sequencing across three matched tissues reveals shared and tissue-specific gene expression and pathway signatures of COPD. Respiratory Research 2019;20. 10.1186/s12931-019-1032-z.

[32] Huang Q, Wang Y, Zhang L, Qian W, Shen S, Wang J, et al. Single-cell transcriptomics highlights immunological dysregulations of monocytes in the pathobiology of COPD. Respiratory Research 2022;23. 10.1186/s12931-022-02293-2.

[33] Ogger PP, Byrne AJ. Macrophage metabolic reprogramming during chronic lung disease. Mucosal Immunology 2021;14:282–95. 10.1038/s41385-020-00356-5.

[34] Fujii W, Kapellos TS, Baßler K, Händler K, Holsten L, Knoll R, et al. Alveolar macrophage transcriptomic profiling in COPD shows major lipid metabolism changes. ERJ Open Research 2021;7:00915–2020. 10.1183/23120541.00915-2020.

