## Supplementary material for "Computational Deconvolution of Cell Type-Specific Gene Expression in COPD and IPF Lungs Reveals Disease Severity Associations": Online Supplemental

**Supplemental Results**

*Difference in signature score across disease*

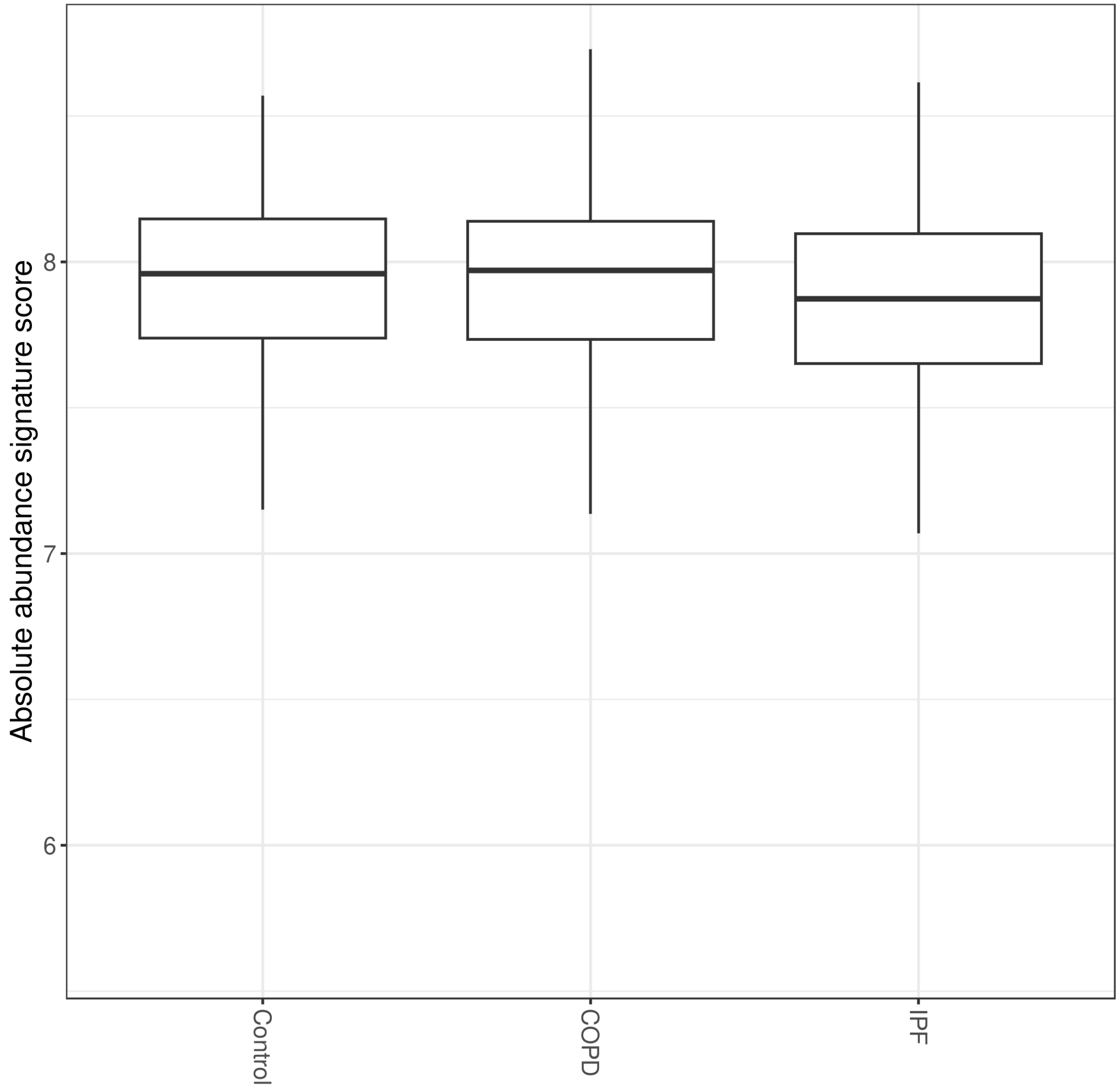


*Figure E1: Boxplots showing CIBERSORTx absolute signature score split by disease status. CIBERSORTx absolute mode was used to calculate the absolute abundance signature score calculated by dividing the median expression level of all genes in the signature matrix by the median expression level of all genes in the sample mixture.*

The absolute score output by CIBERSORTx is estimated by the median expression level of all genes in the signature matrix divided by the median expression level of all genes in the mixture. Therefore, this score reflects whether there was any sample that had cell types in the tissue mixture that was not covered by the cell types represented in the signature matrix. We found that absolute score was lower in IPF samples compared to control (p=0.03) (Figure S1), indicating that IPF tissue may have greater gene expression values from cell types that were not included in our signature matrix. Based on this result, we chose to use the CIBERSORTx absolute abundance score instead of relative proportion for analysis.

*Association between cell type abundance score and GAP score*

Gender (G), age (A), and 2 lung physiology variables (P) (FVC and $D_LCO$) were calculated based on the GAP models [1]. The GAP score uses commonly measured clinical and physiologic variables to predict mortality in patients with IPF. Results are shown in the supplemental Table E4.

*Association between cell type-specific gene expression and disease severity measured by lung function tests*

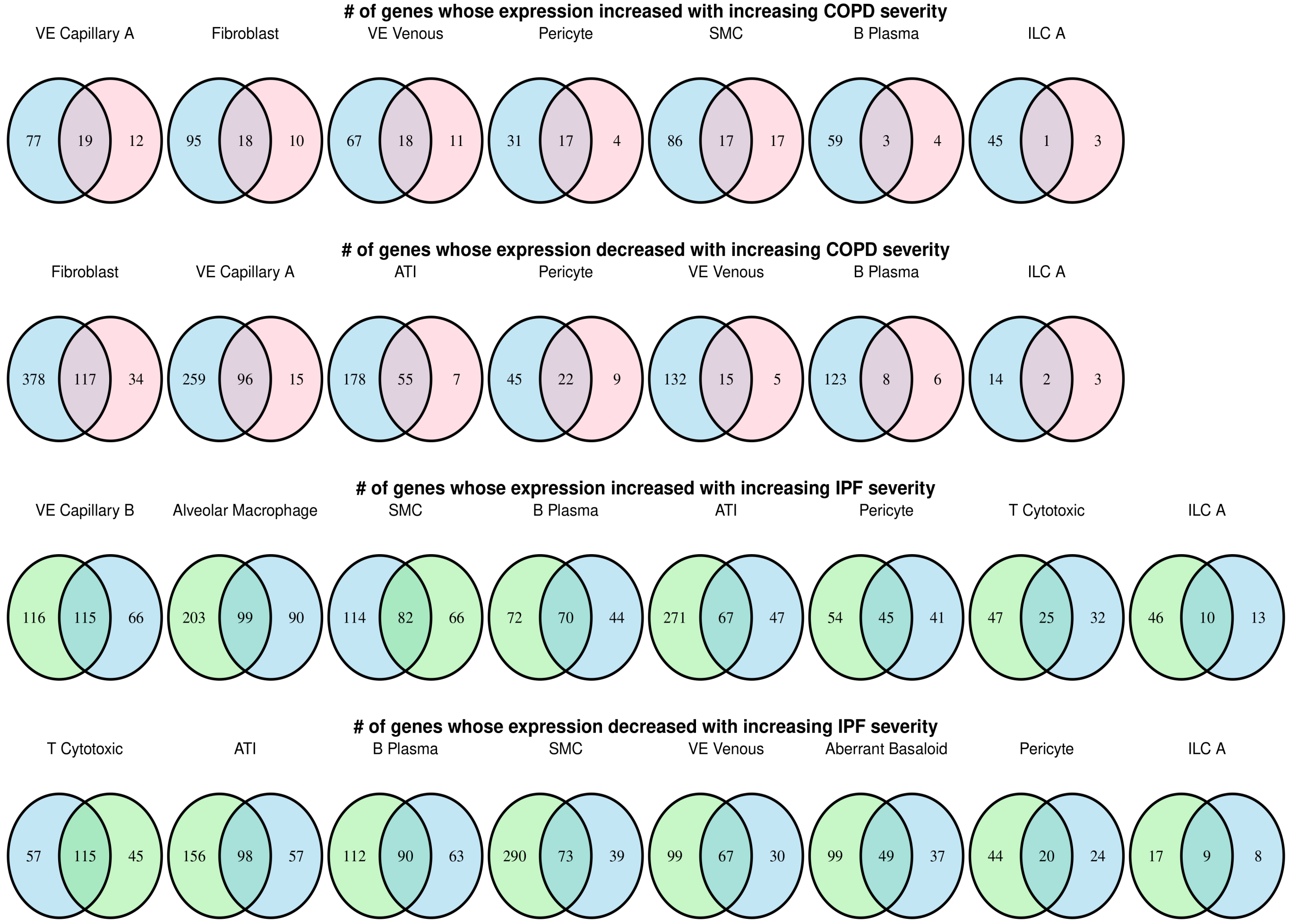


*Figure E2: Venn diagrams showing cell type-specific gene expression associated with disease severity in COPD and IPF lungs. Genes associated with DLCO, FEV1 and FVC are colored blue, pink, and green, respectively. Cell types with the higher number of genes whose expression is associated with disease severity are arranged from left to right.*

**References**

[1] Ley B, Ryerson CJ, Vittinghoff E, Ryu JH, Tomassetti S, Lee JS, et al. A Multidimensional Index and Staging System for Idiopathic Pulmonary Fibrosis. Annals of Internal Medicine 2012;156:684. https://doi.org/10.7326/0003-4819-156-10-201205150-00004.
